# Recruitment, Retention Approaches and Community Engagement in the THRIVE pilot Trial: Lessons Learned from a Food is Medicine Trial

**DOI:** 10.64898/2026.06.12.26355557

**Authors:** Mojisola Olusola-Bello, Elohor Oborevwori, Khadijat Adeleye, Irma Iribe, Adeline Ussani Uva, D’Janee Kyeremeh, Oluwatosin Tomiwa, Janice Dugbartey, Maricielo Leasure, India E Washington, Samuel Gledhill, Chelsea Akubo, Siddharth Dhadi, Mary Alawode, Jennifer Freeman, Kendra Smith, Rose Bernard, Marie Antoinette Davis, Channa Wilson, Anna Maria Izquierdo Porerra, Lisa A. Cooper, Cheryl R. Dennison Himmelfarb, Yvonne Commodore-Mensah, Oluwabunmi Ogungbe, the THRIVE Food Is Medicine Team

## Abstract

**Background:** Recruitment of underrepresented populations, including Black and Hispanic populations, for Food is Medicine (FIM) and cardiovascular trials, may pose significant challenges.

**Methods:** We implemented a multi-component recruitment approach for the THRIVE (AdapTive personalized dietitian coacHing and messaging with pRoduce prescrIptions to improVE healthy dietary behaviors) pilot trial to engage primarily Black and Hispanic adults in a Food is Medicine for hypertension intervention. The recruitment approaches included community engagement at approximately 40 community events (cultural festivals and neighborhood gatherings); partnerships with 8 community and faith-based service hubs and food distribution sites; recruitment through safety net primary care clinics, digital outreach via the study website, and social media campaigns; and direct recruitment at places of worship. We report lessons learned from the community engagement process, recruitment efficiency, representativeness, and retention outcomes.

**Results:** Within 6 months, the enrollment target was exceeded by 40%, with an accrual index of 1.04. Over 1,000 individuals were reached through the direct-to-community engagement process, while faith-based partnerships engaged about 900 adults. There were 2,673 visits to the study webpage, and social media achieved 12,259 impressions with 399 clicks. About 95% of participants resided within 10 miles of the faith-based recruitment sites. Face-to-face engagement at the food distribution sites within faith-based organizations or community service hubs outperformed digital methods. Faith leader endorsements and follow-up in-person meetings (following unsuccessful email outreach) dramatically increased recruitment. Regarding retention, pre-randomization attrition was 6%, and 82% of participants completed the study.

**Conclusion:** Culturally tailored, community-engaged recruitment grounded in faith-based and local community partnerships, was highly effective in engaging Black and Hispanic populations in this FIM cardiovascular trial. This provides a replicable model for implementing equitable and sustainable cardiovascular health interventions.

## BACKGROUND

Cardiovascular disease(CVD) is the leading cause of mortality in the United States.^1^ There are substantial cardiovascular health inequities experienced by racial and ethnic minoritized groups.^2^ For instance, Black adults face a higher age-adjusted CVD mortality rate compared to their White counterparts.^3^ Similarly, Hispanic adults have a higher prevalence of CVD risk factors.^4^ Food is Medicine (FIM) interventions, which may include produce prescriptions, medically tailored groceries, and medically tailored meals with nutritional support, nested within healthcare delivery, have emerged as promising approaches for cardiovascular disease prevention and management.^5–8^ However, the recruitment of people who are most affected by nutrition insecurity and diet-related chronic conditions for FIM trials presents challenges^9^ that differ substantially from those of other trials.

Black and Hispanic individuals are overrepresented in communities facing significant household food insecurity,^10^ leading to poor dietary quality and disproportionately high rates of CVD and other diet-related chronic conditions.^11–13^ These populations are also more likely to live in healthy food priority areas () for FIM studies, to have a history of historical segregation, and to experience underinvestment in these neighborhoods.^14,15^ Despite this, participation by minoritized populations in clinical trials, where they are likely to benefit most, remains dismal.^5^ Some reasons for this include inadequate contextually tailored recruitment strategies, the uncritical transplantation of conventional recruitment models into community-engaged contexts where they were never designed to function, the absence of explicit enrollment goals, and historical and ongoing research misconduct that negatively impact the minoritized community.^16–19^

Successful participant recruitment is widely recognized as one of the most challenging aspects of clinical research, with implications for study validity, generalizability, and eventual translation to clinical practice.^20,21^ Recruitment challenges may show up in identifying potential participants, engaging referral partners, and retaining participants in the study.^22^ This is particularly pronounced in community-based interventions among minoritized populations, where passive recruitment methods are insufficient due to mistrust of research institutions, healthcare professionals, and researchers; socioeconomic and logistical challenges; language and cultural barriers; lack of awareness about ongoing studies and trials; external influences; perceived bias; and competing life priorities.^23^ This necessitates culturally sensitive, community-informed recruitment strategies that address both clinical eligibility and practical considerations, particularly in FIM trials, such as meal acceptance, delivery logistics, and the sustained implementation of dietary changes over time.^24^

Despite the growing implementation of FIM trials and programs across healthcare systems, clear documentation of recruitment strategies for these interventions is limited. The rapid expansion of FIM programs in clinical settings fundamentally shows the need for evidence-based recruitment strategies to support robust research and implementation. In this paper, we describe the recruitment methods used to recruit adults with hypertension living in HFPAs areas into the THRIVE FIM pilot trial. We also provide lessons learned and a road map for recruiting and retaining community-dwelling participants in future FIM studies.

## STUDY APPROACH

### Study Setting and Context

The THRIVE (AdapTive personalized dietitian coacHing and messaging with pRoduce prescrIptions to improVE healthy dietary behaviors) pilot trial (NCT06257550)^25^ was a two-arm randomized pilot trial that assessed the feasibility of a culturally tailored FIM intervention. The intervention combines produce prescriptions, personalized dietitian coaching, adaptive messaging, and linkages to social resources to improve dietary behaviors among participants with hypertension. THRIVE, funded by the American Heart Association’s Health Care by Food initiative, was conceptualized within the broader FIM framework, which recognizes food interventions as essential components of healthcare delivery and chronic disease management.^26^ This framework positions access to nutritious food as a social and health service and a fundamental element of medical care.^26^

#### Geographic Focus

The THRIVE study was conducted in Montgomery County, Maryland, a region characterized by stark socioeconomic contrasts (**Figure 1**). Despite its reputation as one of the wealthiest counties in the United States, approximately 45% of Montgomery County households earn less than the self-sufficiency standard, forcing difficult choices among housing, childcare, healthcare, and healthy eating.^27,28^ The county’s population is notably diverse, with one-third of residents being recent immigrants, many of whom face additional barriers to accessing federal benefits and healthcare services.^29^ The study focused on Healthy Food Availability Index scores, defined as a composite index incorporating food availability, economic indicators, and accessibility metrics.^30^ These areas are characterized by low HFPAs and significant economic and transportation barriers to accessing nutritious food. Clusters of HFPAs are particularly concentrated in the eastern and southern regions of the county, areas that also showed a higher prevalence of cardiovascular disease and related risk factors.

**Figure 1.**
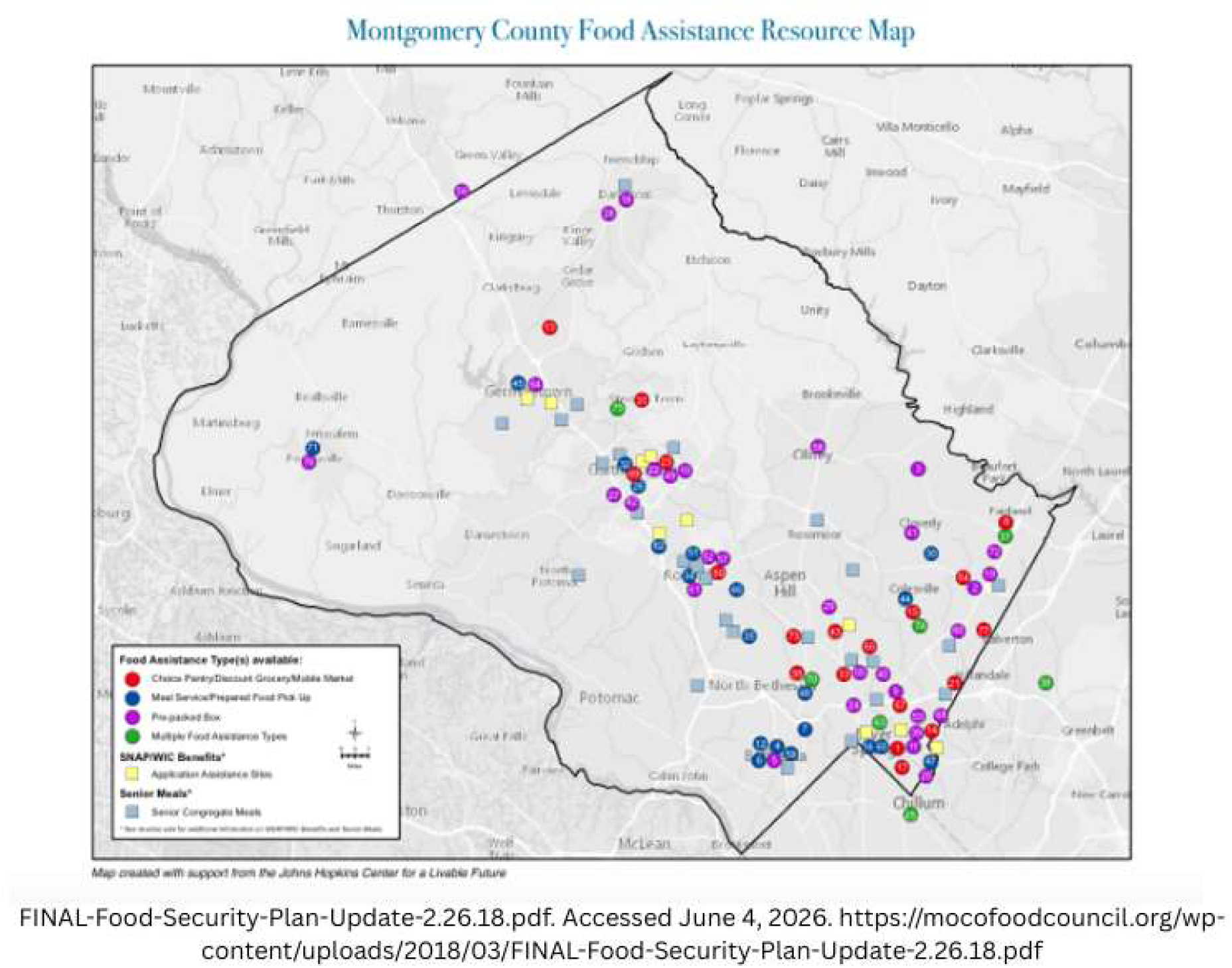
Montgomery County Healthy Food Priority Areas and THRIVE Study Sites.

#### Food Security Landscape

According to a 2025 Hunger Report by the Capital Area Food Bank, 35% of Montgomery County residents are experiencing food insecurity.^31^ The county has the highest rate of eligible but unenrolled beneficiaries of the Supplemental Nutrition Assistance Program in Maryland.^28^ Approximately 64% of eligible residents do not receive these benefits, highlighting significant gaps between resource availability and utilization.^27^

#### Site Selection Considerations

The selection of engagement and recruitment sites was based on our team’s consideration of geographic and demographic factors as well as local food economics^28,31^ Through partnership with Community FarmShare,^32^ a local food systems initiative, the study sites were strategically positioned to strengthen connections between local sustainable farmers and communities experiencing food insecurity. Community FarmShare’s existing relationships with more than a dozen farms in Montgomery County enabled the study to support the local agricultural economy and ensure that high-quality, fresh produce is provided to participants. Priority was given to locations within or adjacent to identified HFPAs that could serve as established community gathering points and nodes (e.g., Montgomery County consolidated community service hubs) in a sustainable local food distribution network.

Healthcare access in the target communities reflects similar patterns of disparity, with significant barriers to both preventive and chronic disease care. Transportation limitations, financial constraints, and cultural and linguistic obstacles create complex challenges for residents seeking healthcare services. Our study partnered with 2 health systems that have mobile primary care models to support residents by addressing these barriers. Care for Your Health (C4YH)^33^ is a nonprofit health system that delivers inclusive, multicultural healthcare. C4YH’s innovative “pop-up” mobile clinic model was key to reaching underserved neighborhoods, providing a unique recruitment approach and a means of delivering healthcare, addressing common barriers to access. Mobile Medical Inc.^34^ was another nonprofit healthcare organization that provides primary healthcare and proactive management of chronic diseases to insured, uninsured, low-income, and Medicare– and Medicaid-enrolled residents of Montgomery County.

## RECRUITMENT APPROACH

We employed both active outreach and passive recruitment.^35,36^ For the human-centered design phase of the trial, we engaged community residents and key partners in human-centered design sessions to incorporate diverse perspectives.^37^

### Active Outreach Strategies

Active outreach was our primary recruitment strategy, particularly through multiple community screening events. These events were strategically organized in locations already frequented by our target population, minimizing transportation barriers, and were often integrated into existing, well-known community events. We also leveraged community service hubs, multipurpose centers run by community-based organizations, and the Department of Health and Human Services in Montgomery County^38^ collaborating with other nonprofit partners to serve Montgomery County residents who need food and other resources. The food distribution services through the service hubs became strategic recruitment locations, given their alignment with the study’s focus on nutritional interventions. Identifying these areas as places where HFPA members already gather, our team formed partnerships with the hubs and established a regular presence at these sites (Figure 2). Our team also established a regular presence at other local food pantries, engaging directly with individuals already seeking food resources, including approaching participants in lines of parked cars as they waited for the distribution to begin. We provided free health screenings and health promotion activities and informed potential participants about the study upon interest.

**Figure 2.**
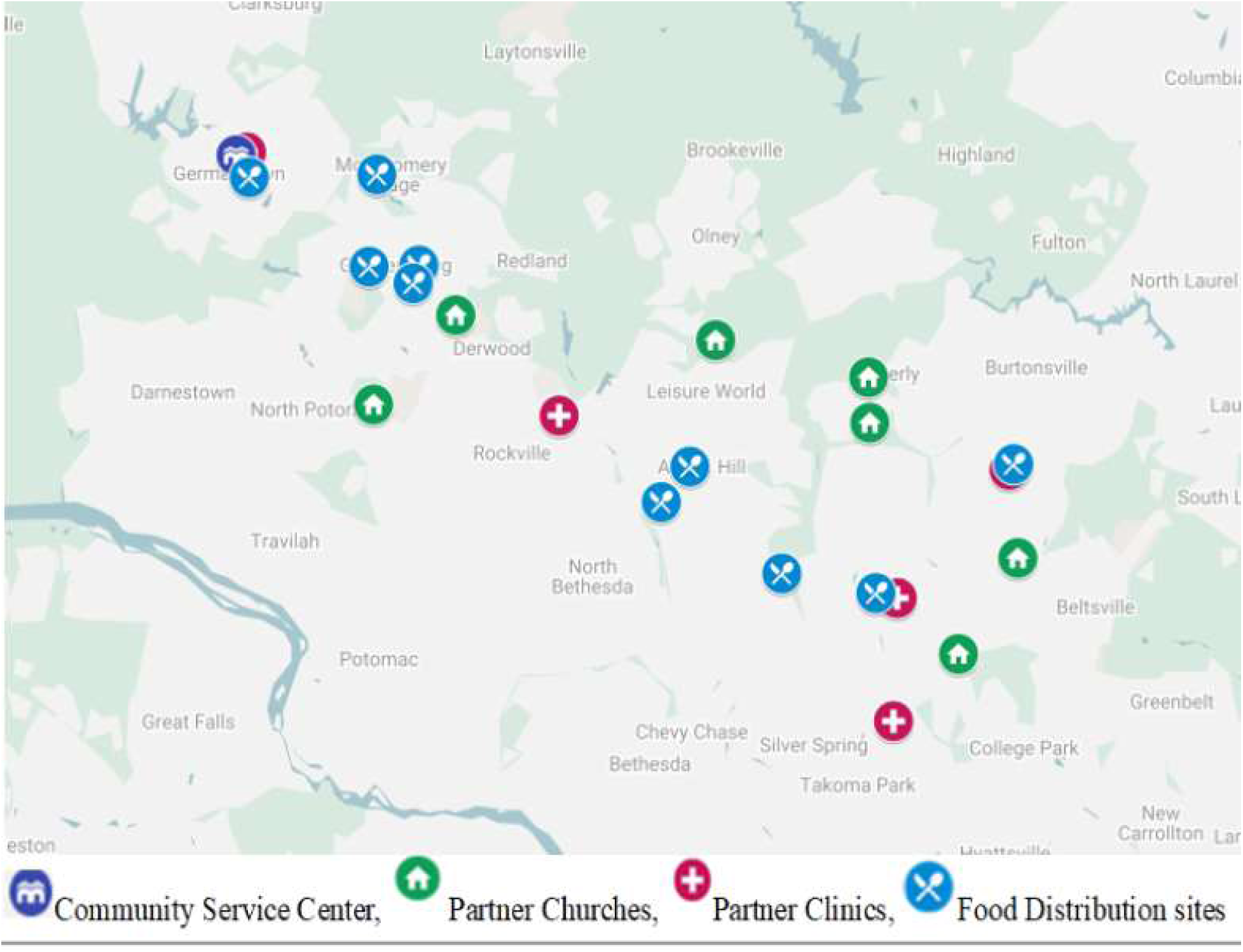
Map of the Churches, Clinics, and Food Distribution Sites in the THRIVE Trial.

Through our partnerships with C4YH and Mobile Med Inc. clinics, the clinic physicians and staff distributed the study flyer to their patients. These potential participants were encouraged to scan the QR code on the flyer, which linked to the study interest form, if they were interested in participating.

We identified church leaders as key gatekeepers within faith-based organizations. After establishing relationships and securing their support, we partnered with them to engage their congregations. With their buy-in, we hosted recruitment events immediately following church services, leveraging their influence and trust within the community to enhance participation.^39^ These partnerships enabled community members who might otherwise remain disconnected from research opportunities. As shown on the map, we established partnerships with 7 churches across various locations (**Supplemental Figure 1**). The communications team from one of our partnering churches invited our team to record a recruitment video, which was played during announcements during church services and displayed on the study website.^40^

Community celebrations and gatherings were additional recruitment venues in more informal settings. The research team participated in community picnics, cultural festivals such as the Ewe Festival (festival for Ghanaian immigrants from the Ewe tribe), and neighborhood events, with the goal of continuously establishing a visible presence at these gatherings. These allowed for more conversational introductions to the study, often facilitated by community organization leaders who shared cultural and linguistic backgrounds with attendees.

### Passive Recruitment Approaches

Complementing the active strategies, we implemented several passive recruitment approaches designed to extend reach beyond in-person encounters. We conducted social media advertising on Facebook and Instagram using the detailed targeting feature to ensure the ads are shown to residents of the specified zip codes with tailored content.^41,42^ This social media strategy included both English– and Spanish-language content, with culturally relevant imagery and messaging.

The research team regularly posted flyers on community boards at grocery stores, pharmacies, and community centers. These materials used clear, accessible language in both English and Spanish, with prominent contact information and QR codes that linked to the study recruitment page. Word-of-mouth referrals were also used as the study recruitment progressed. Early participants often referred family members and friends, including church members, which created an organic community-recruitment network.

### Digital Trials Platform

We leveraged digital recruitment strategies through partnering with Studypages,^43^ a remote trials platform, to support our recruitment efforts. Through this partnership, we launched a dedicated recruitment webpage^40^ which displayed eligibility requirements, participation details, and contact options, and was optimized for accessibility and available in English and Spanish. The page included an embedded sign-up form, allowing interested individuals to register easily. Submitted information was securely routed to the study email for timely follow-up. Studypages^40^ also served as our primary recruitment management system.

### Recruitment Incentives

Participants were informed in advance about the incentives provided for their participation. Those who participated in the human-centered design sessions received $300 in total compensation for attending all 3 co-design sessions (each session lasted 2 hours).^37^ During the intervention phase, participants were informed that they would receive $35 Tango e-gift cards upon completion of each data-collection visit and the qualitative interview, for a total of $140. In addition, participants would receive $30 worth of fresh produce each week throughout the study.

One of the key partner churches,^44^which regularly distributed millions of pounds of food and other essential items, provided gift cards to encourage participation in the initial screening process at their location.

At enrollment, participants received measuring utensils and mesh produce bags and were informed that at the 12-week mark, they would receive a kitchen scale and a reusable water bottle. In addition to these, an interactive prize giveaway was held at 12-week follow-up visits, allowing participants to spin a wheel for a chance to win an air fryer, a smoothie blender, or a coffee maker. Upon completing their 24-week study, participants received thank-you letters and vegetable seed packets to help maintain healthy eating habits at home.

## SCREENING AND ENROLLMENT PROCESS

Two main steps (initial pre-screening and eligibility screening) were used to determine participants’ eligibility.

### Initial Pre-Screening

The initial survey screening process involved a self-reported web-based pre-screening form, which could be either self-administered through our Studypages website^40^ or assisted by members of the THRIVE team. This screening form was on the Studypages platform,^43^ the primary recruitment management system for the study. At the end of the pre-screening form, the system automatically identified ineligibility based on the respondent’s responses and displayed a message explaining the reasons for ineligibility. The Studypages platform categorized individuals into 2 primary groups: sign-ups and participants; each subdivided into 2 statuses. Active sign-ups were individuals interested in study participation who had not yet been screened by the team. In contrast, completed sign-ups included individuals who were no longer interested or deemed ineligible. Participants were considered active after they had been screened and enrolled, and inactive if they withdrew or lost to follow-up.

### Eligibility Screening

Participants who were eligible after the pre-screening process were invited to an in-person eligibility screening visit at the closest, most convenient community site for them. This included blood pressure screening, measured with automated monitors after participants had rested for at least 5 minutes, in accordance with AHA guidelines for blood pressure measurement.^45^ Point-of-care hemoglobin A1c (HbA1c) testing was performed using finger-prick blood samples to assess diabetes status. Physical measurements, including weight and height, were taken. Participants were also asked about any current health conditions, including gastrointestinal and diet-related conditions, as well as food intolerances, that might make them unsuitable for the study. These were conducted in a conversational format using a standardized questionnaire. Additionally, all participants, regardless of eligibility, were asked about their social and healthcare access needs and were provided with information to connect them with these resources.

### Consent and Enrollment Procedures

When possible, the in-person eligibility screening was combined with the enrollment visits. The consent process was conducted with eligible participants. The study surveys were then administered; if not on-site, a phone option was offered and was scheduled at times convenient for participants, including evenings and weekends. Thus, survey administration occurred primarily via telephone interviews or in-person meetings, often after a church service or clinic visit, to accommodate participants’ limited time. To complete the enrollment process, venous blood samples were collected by our C4YH clinic partners to assess the study panel of cardiometabolic and kidney-function laboratory tests. Participants also completed a 24-hour dietary recall administered by dietitian interviewers. Participants received reminder calls or text messages for follow-up assessments (**Figure 3**).

**Figure 3:**
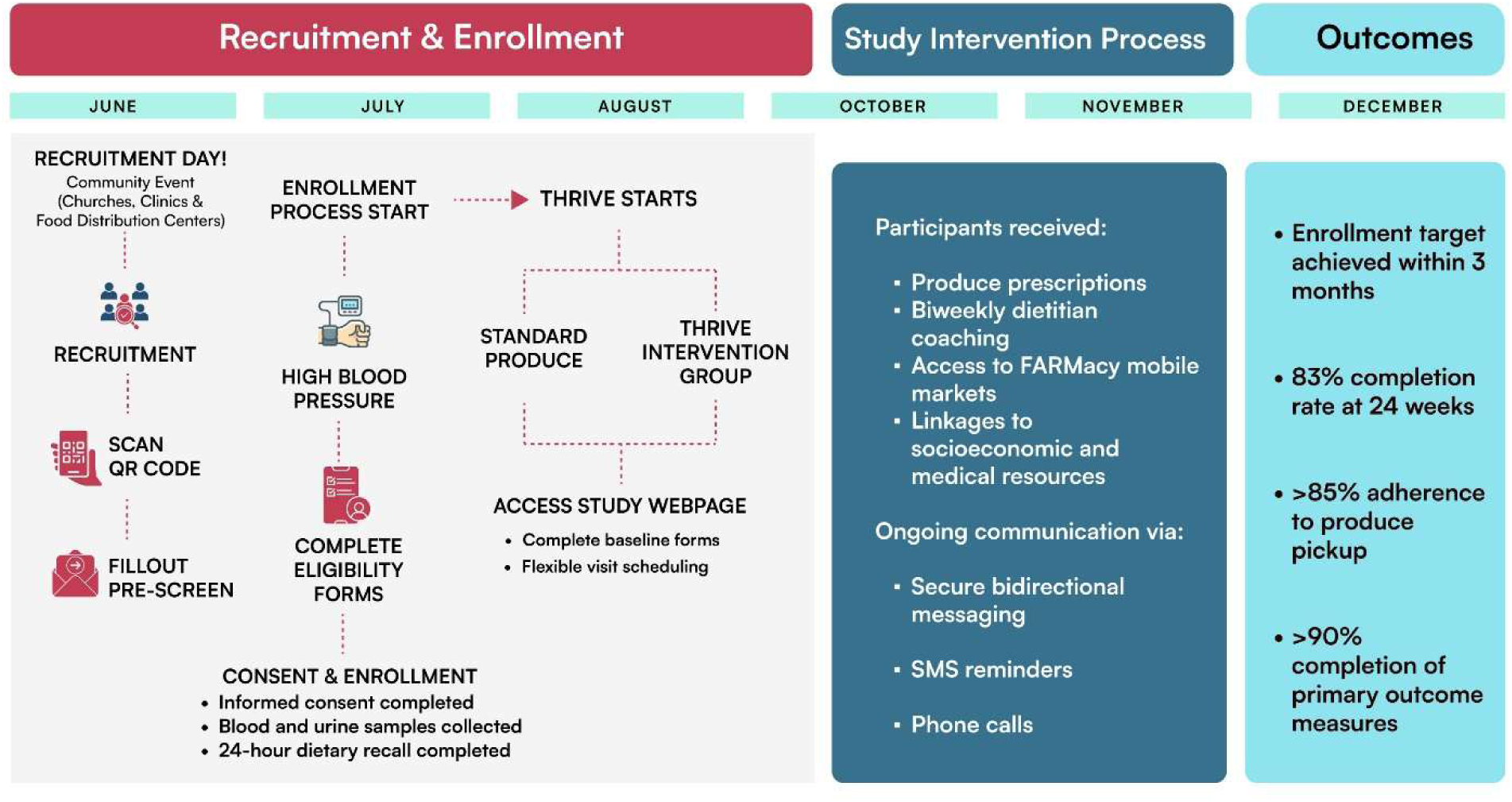
THRIVE Recruitment process.

**Figure 4:**
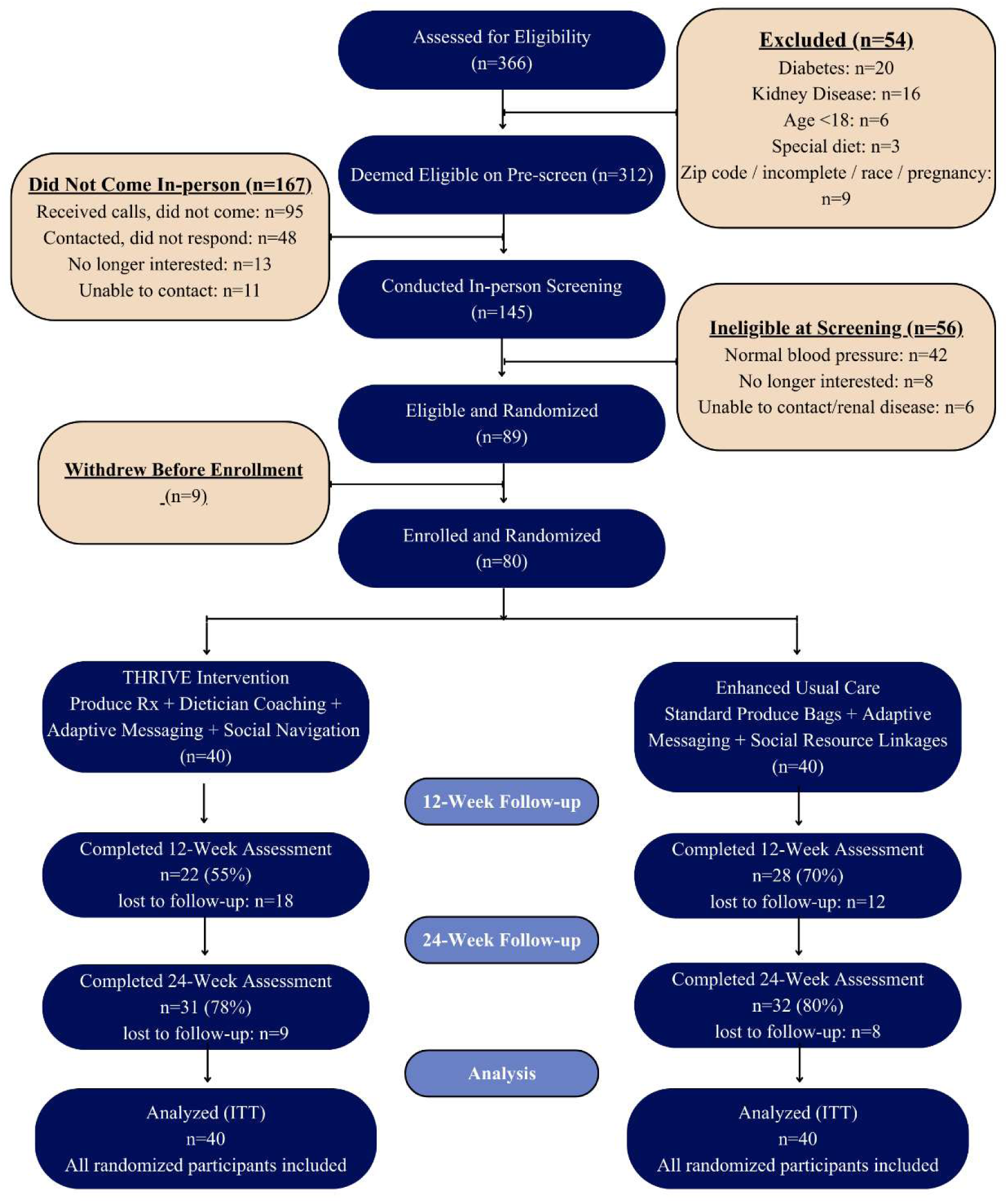
Consort flow diagram.

Communication with participants was conducted through the Studypages platform, which included secure messaging, SMS, and phone calls. To access the secure messages, participants received a text notification containing a secure access link that required verification via an SMS code. In contrast, direct SMS messages were delivered to participants’ devices with no additional steps. The team adhered to the protocol for choosing between secure messages and SMS, often based on message content. The platform supports bidirectional messaging, allowing participants to respond to regular SMS messages as usual, while secure messages require verification before response.

## LESSONS LEARNED

As a result of implementing the recruitment strategies detailed above, the study met its enrollment target within 3 months (June to August) and initiated a second phase of recruitment between November and December in response to requests from community partners (**Figure 3**). There was remarkable efficiency during the recruitment phase, with participants typically proceeding from initial contact to screening within 1 to 2 weeks. Many individuals were screened on the same day they learned about the research. The cornerstone of recruitment success was the active community engagement strategy, particularly recruitment conducted at places of worship, which proved far superior to other methods in both reach and conversion rates.

During the first and second recruitment periods (June-August 2024 and November-December 2024), the research team attended approximately 40 community events across Montgomery County, including community picnics, cultural festivals (notably the Ewe Festival held in Gaithersburg, Buganda Day in Silver Spring, etc.), and neighborhood gatherings (such as the June 15 Humanitarian Resource Event at the East County Regional Service Center to welcome new immigrants) at various community centers and parks. These events collectively attracted hundreds of attendees, providing extensive opportunities for direct community engagement.

In addition, the research team formed partnerships with approximately 8 food distribution sites in churches across Montgomery County, establishing a weekly or biweekly presence throughout the recruitment period. With pastoral endorsement, recruitment events were held immediately after church services, reaching hundreds of congregants. The QR code for the sign-up sheets was made available at all community events and church services, yielding approximately 355 on-site sign-ups across all venues. The participant onboarding process was successfully implemented through the digital enrollment system.

The study website received 2,673 visits during the active recruitment period, with an average engagement time per active user of 1 minute 11 seconds. The views per active user were 4.11, and the number of active users was 252. Additionally, targeted social media (Facebook and Instagram)^41,42^ campaigns reached 12,259 unique individuals. There were 18,736 ad impressions, generating 399 unique clicks to the study’s Studypages website. The social media strategy effectively targeted key demographics, with the highest engagement among college-aged individuals (18-24 years: 5,050 impressions), young adults (25-34 years: 3,869 impressions), and seniors (65+ years: 3,954 impressions).

### Enrollment and Retention Feasibility

The study successfully enrolled 80 participants, with an accrual index of 1.05 (calculated as the ratio of observed to expected accrual at monthly intervals throughout the recruitment period). Participants were randomized into 2 groups: 40 in the intervention group and 40 in the control group. Among the 80 enrolled participants, 9 were lost to follow-up (unable to be contacted), 4 withdrew, and 2 experienced health issues post-randomization, resulting in an overall completion rate of 78% (**Figure 4).** Adherence to the produced prescription pickup was markedly high, exceeding 85%.

**Figure 5:**
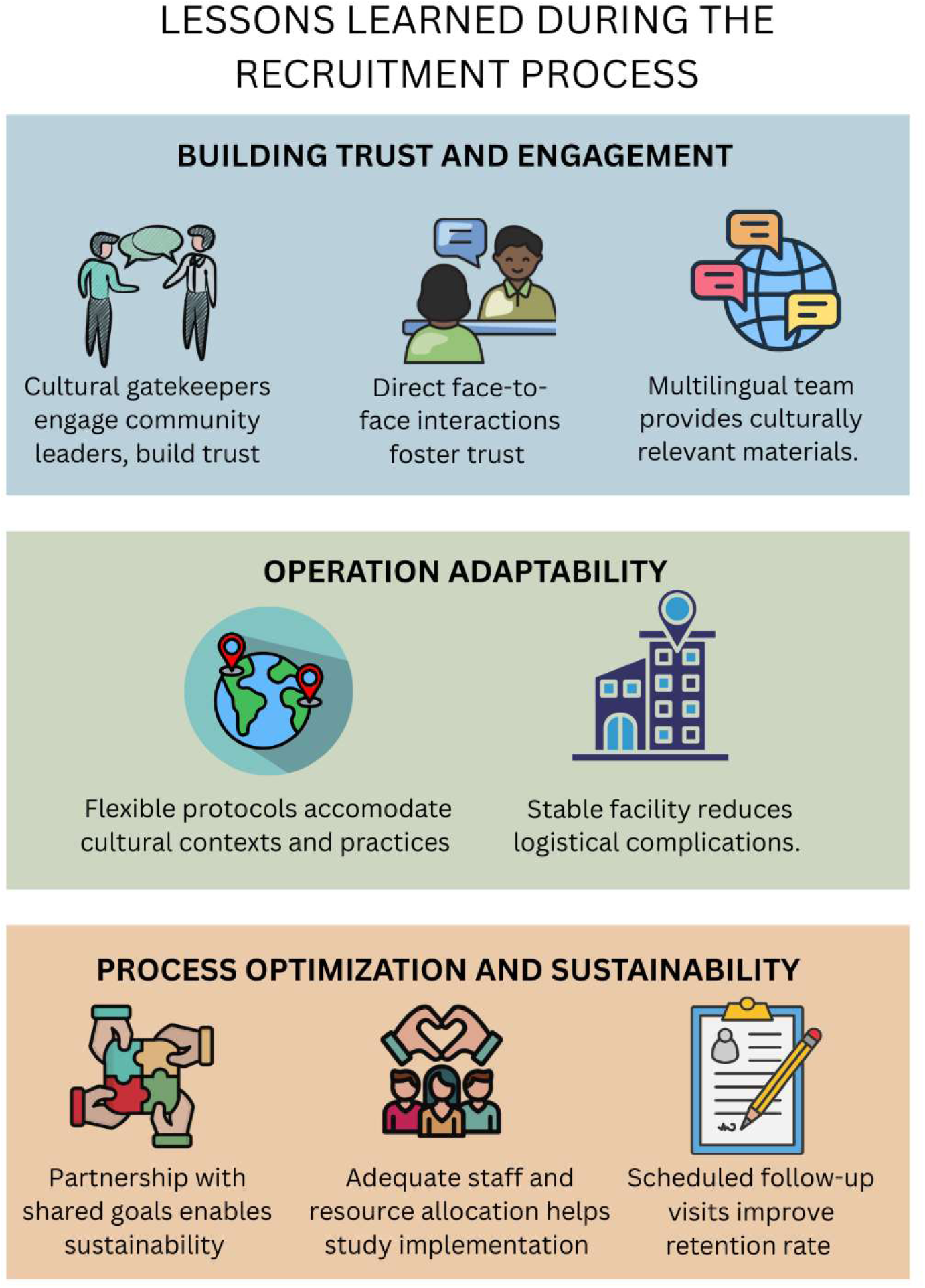
Lesson Learned During the Recruitment Process.

### Lessons Learned from the Recruitment and Retention Process

#### Importance and Role of Community Leaders

In our recruitment strategy within HFPAs, we recognized that genuine engagement with community gatekeepers is essential to successful participant enrollment, particularly in communities with a history of mistrust of research. Early in the process, we attempted to recruit participants by distributing flyers but received a minimal response and observed that participants appeared hesitant to approach us and often avoided eye contact.

The breakthrough came when a research team member who attended one of the churches we were engaging with arranged a face-to-face meeting with the pastor. Initially, our emails to church leaders had gone unanswered for weeks. By prioritizing listening over presenting what we intended to achieve, we learned what mattered most to this community leader and his community members. We shared feedback and experiences from our human-centered design phase and demonstrated our commitment to mutual benefit. We explained how community input had already shaped our program design, and we outlined our plans to share results and sustain program elements beyond the research period. Subsequently, we secured his endorsement; he personally introduced us during the Sunday service and appeared in a video announcement shown at multiple churches. Within 2 weeks, our recruitment numbers increased dramatically. This showed that engagement requires more than seeking permission to recruit; it also requires genuine partnership, demonstrated commitment to community benefit, acknowledgment of historical harms, and transparency about research goals and community returns.

#### Recruitment Efficiency and Quality Over Quantity

Another important lesson was recognizing the distinction between recruitment volume and recruitment quality, especially as potential participants face multiple competing demands and limited capacity for sustained engagement with programs. Our teams’ participation in large-scale community events, such as free product giveaways and promotional activities, generated numerous sign-ups and appeared initially successful in terms of numbers. However, these participants had dramatically lower retention rates than those recruited through our church-based partnerships.

The differentiating factor was community embeddedness. Participants recruited at large promotional events often resided outside our areas of focus and had no affiliation with our partner churches or community organizations. Without the social accountability of community involvement or the practical convenience of geographic proximity, these individuals rarely progressed beyond the initial visits. This pattern persisted despite our efforts to retain them, including numerous follow-up communications, transportation assistance, and flexible remote-visit options via video or phone.

We learned that recruitment strategies that yielded high enrollment numbers are not necessarily the most efficient or cost-effective if they fail to attract participants with characteristics that predict sustained engagement. Individuals embedded within the community networks we partnered with, who saw familiar faces at study visits, whose participation was known to their pastor or community leaders, and who lived near intervention delivery sites, had built-in accountability relationships and fewer logistical barriers, which facilitated long-term retention.

#### Influence of Face-to-Face Interactions

A key lesson from our recruitment efforts was the clear superiority of face-to-face interaction in trusted community spaces over passive, virtual, or digital recruitment methods. First, in-person interaction offered invaluable opportunities to build trust and reduce barriers. When we met potential participants face-to-face, we could address their questions, concerns, and skepticism in real time, often converting hesitation into enrollment in a single conversation. Concerns about research motives, time commitment, confidentiality, or study procedures could be addressed with nuance and compassion that were impossible to convey through written materials or social media posts. Second, physical presence in community spaces demonstrated respect and cultural humility in ways that remote recruitment could not. Being at spaces where community members already gathered, rather than expecting them to seek us out or respond to impersonal advertisements, signaled that we valued their time and were willing to meet them where they were.

Additionally, to sustain their engagement, we assigned each participant a primary study contact who maintained regular communication throughout the study period, not only for scheduled assessments but also for check-ins and support. This consistency fostered trust and continuity in relationships. Participants said they appreciated that the team members knew their name, remembered their circumstances, and genuinely cared about their well-being beyond data collection. Building on the trust established during initial face-to-face recruitment encounters, these ongoing personal connections sustained engagement over time.

#### Supportive Multilingual Team

One of our most important lessons was recognizing that language accommodation was a fundamental pillar of ethical research practice, equitable access, and authentic community engagement. Initially, we may have viewed multilingual services as a “nice to have” feature that would broaden our eligible participant pool. However, we learned that language accessibility was central to nearly every aspect of successful recruitment, retention, and intervention delivery. Our experience taught us that offering study participation in multiple languages, with team members conducting screening and obtaining informed consent in English, Spanish, Creole, French, and Twi, was essential.

Importantly, we learned that language capacity is an equity issue in research, determining who can participate in research and access its potential benefits. We built a multilingual team, ensuring that immigrant communities and linguistic minorities could participate in the study, access that they would have been denied if English proficiency had been an implicit or explicit requirement for participation.

The benefits of this investment in a multilingual team became evident in our recruitment and retention metrics. Community members who could participate in their native language enrolled at notably higher rates than our initial approach without native language speakers when approached, demonstrated better retention throughout the study, provided higher-quality data (such as more complete dietary recalls, more detailed barrier descriptions, and more honest feedback during the qualitative interviews), showed greater engagement with the intervention, and reported a high satisfaction with the program.

#### Balancing Protocol Requirements and Cultural Tailoring

One of our most challenging lessons involved navigating the tension between standardized research protocols required for scientific validity and the cultural adaptability necessary for meaningful community engagement and sustained participation. We initially approached the study with strict adherence to a predetermined protocol that was a better fit for rigid, clinic-only trials.

This approach created significant implementation challenges that threatened both recruitment and retention in HFPAs. Community members, partner organizations, and our front-line staff identified numerous aspects of our protocols that conflicted with cultural practices, failed to accommodate participants, imposed unnecessary burdens without clear benefits, or created barriers that disproportionately excluded the most vulnerable community members we aimed to engage. We recognized that scientific rigor and cultural adaptability can be interdependent. Research that fails to engage and retain participants, particularly those most affected by the health disparities we aimed to address, produces no valid findings, regardless of protocol perfection. This realization led us to develop an approach where we distinguished between core intervention components essential to the research question (which required consistency to allow valid comparisons and causal inference) and implementation details that could be flexibly adapted to local context (which benefited from community-driven modification to enhance engagement, cultural appropriateness, and real-world applicability).

#### Designated Space for Data Collection in the Community

Our team intended to secure designated, stable study locations within the community from the project’s inception. Having consistent, reliable facilities was instrumental in keeping the study on schedule. This stability helped participants become familiar with the study sites, reduced logistical complications, and enhanced the research team’s professional credibility. Future studies should prioritize early negotiation and formal agreements for dedicated research space in the community, as this element significantly affects both recruitment efficiency and participant retention by providing a consistent, accessible, and welcoming environment for study activities.

#### Community Partners’ Engagement and Alignment of Priorities

Engaging community partners for recruitment can be challenging, particularly when organizational priorities diverge from study timelines. We learned that scheduled screening events were vulnerable to postponement due to competing events at recruitment sites and that assumptions about access permissions were often inaccurate. For example, we encountered situations in which service center leadership had not communicated permissions to frontline staff, preventing us from approaching potential participants at community hub distribution sites. This experience taught us the importance of establishing clear, transparent, and documented communication processes with all levels of partner organizations, from leadership to operational staff. We learned to formalize agreements in writing, establish regular communication checkpoints, and build buffer time into recruitment schedules to accommodate partner organizations’ other activities.

#### Staffing and Resource Allocation

We learned that hiring adequate research staff was essential for efficient study implementation, and that budget constraints in this area created cascading challenges throughout the project. Our heavy reliance on student volunteers was insufficient to achieve rapid progress and maintain consistent study momentum. Some volunteers, despite their dedication, could not provide the needed continuity, accountability, and commitment. This experience highlighted the importance of realistic budget planning that prioritizes adequate staffing levels, as understaffing ultimately compromises recruitment rates, data quality, and retention efforts. Future research should account for sufficient personnel costs in grant applications and seek supplementary funding streams specifically for staffing needs. Additionally, we learned that a hybrid model combining a core team of paid staff with strategic volunteer support for specific tasks can optimize efficiency and community engagement while remaining budget-feasible.

#### Proactive Participant Contact and Follow-Up Reminders

We learned that establishing a protocol for contacting participants before their scheduled follow-up visits was essential for retention and logistical planning. Rather than waiting for participants to arrive on their scheduled dates, we made reminder calls or sent messages several days before their appointments. This was highly effective in improving attendance rates by enabling participants to reschedule conflicts early, ensuring alignment between their availability and our study schedule. Beyond improving show rates, contacting them in advance helped us identify participants who were becoming disengaged or facing barriers to continued participation. Recognizing these patterns early enabled us to implement retention strategies, such as providing transportation assistance, offering flexible scheduling, or addressing specific concerns, before participants were lost to follow-up.

Having a sense of the expected attendance also enabled our research team to allocate staff and resources more efficiently, avoiding wasted preparation for no-show appointments and allowing us to adjust daily schedules to maximize productivity. This proactive communication approach was very helpful in shifting our follow-up process from reactive to strategic, thereby improving both retention rates and operational efficiency.

#### Use of Digital Tools

Despite significant investment in social media campaigns, online advertising, and website development, digital-only recruitment approaches yielded minimal enrollment. We learned that while participants may own smartphones and use social media, they are unlikely to discover, trust, or respond to program information from unfamiliar organizations that appear digitally without a prior connection.

Our website functioned most effectively as a supporting resource for in-person recruitment rather than as a stand-alone enrollment generator. Analytics revealed that successful enrollees typically spent minutes on the website, viewing just the homepage and eligibility information. We learned to reframe these brief visits as indicators of efficient information transfer—participants were using the website to quickly verify details shared by trusted community sources (pastors, food pantry staff, community health workers), not for extended exploration.

Based on this insight, we redesigned our website as a streamlined “digital information hub,” prioritizing: front-loaded essential information on the homepage (eligibility, benefits, locations, contact methods); prominent click-to-call phone numbers and text message options; mobile-first design for smartphone access; immediately accessible multilingual content; and minimal text using bullet points and simple language.

#### The Research Team Becoming a Part of the Community

As the study team, we initially perceived ourselves as external to the community. During implementation, we learned that we became part of the community. Participants invited us to family events; church leaders asked us to participate in community initiatives beyond the scope of our study; and community members began to see us as a resource for health questions unrelated to our research. This was part of the trust-building process, such that our engagement with the community was not extractive. We learned to honor the relationships we built by continuing engagement beyond data-collection periods, sharing resources even when they were not directly related to our study, and maintaining relationships after the research concluded. We continue to receive invitations to support community health fairs with screening and health promotion activities, and we honor these invitations as much as we can. Our key takeaway was that community-engaged research could modify the role of researchers from mere external observers to trusted partners with the community, with continuous relational obligations.

### Community Dissemination of Findings

In keeping with our commitment to community-engaged research, our team prioritized returning results to the communities that made this work possible. On April 30, 2026, we partnered with the Maryland chapter of the Preventive Cardiovascular Nurses Association (PCNA) to present our pilot findings at its annual Spring community education program, attended by approximately 49 community, academic, clinical, and public-sector partners. A moderated panel, featuring three study participants, a THRIVE-registered dietitian, the Executive Director of Community FarmShare (our produce partner), and a representative from the Care for Your Health clinic, facilitated rich dialogue about their experiences as participants in the study or partners with our team, the study’s impact, and how they have sustained the intervention components as practical implications for cardiovascular health promotion. This exemplifies the bidirectional nature of community-engaged research in a way that is not extractive. Healthcare professionals who were in attendance engaged directly with participants and our team members, asking how to replicate nutrition-focused community approaches in their own settings. Attendees left with bilingual result summaries and expressed strong interest in future dissemination events, reflecting both the reach and resonance of the THRIVE model beyond the trial itself (**Supplemental Figure 1**).

## CONCLUSION

Recruiting and retaining Black and Hispanic adults in trials requires a different paradigm than the current model. In our experience, the THRIVE pilot trial demonstrates that this is achievable and efficient when recruitment is grounded in genuine community partnership rather than transactional outreach. Our processes were successful because we employed a multifaceted approach that positioned the research team as trusted community members rather than as external investigators seeking participants. As FIM programs expand across healthcare systems nationally, the demand for evidence-based, community-centered recruitment models will grow. The strategies documented here offer a replicable framework for investigators seeking to engage populations most affected by nutrition insecurity and cardiovascular disease, the very populations whose inclusion is essential to ensure that FIM research produces findings that are both scientifically valid and generalizable.

## SOURCES OF FUNDING

The THRIVE Food Is Medicine Study is funded by the American Heart Association Health Care by Food Initiative (grant number 24FIM1264121). Dr. Ogungbe’s research is supported by the American Heart Association Career Development Award (25CDA1453103), and the American Heart Association Implementation Science Award (25ISA1453143), and the American Heart Association Healthcare x Food Award (25FIMPG1466841). Dr. Ogungbe is also partially supported by the National Institute of Diabetes and Digestive and Kidney Diseases (NIDDK), the Office of Disease Prevention (ODP), the Office of Nutrition Research (ONR), the Chief Officer for Scientific Workforce Diversity (COSWD), and the Office of Behavioral and Social Sciences Research (OBSSR) of the National Institutes of Health under award number U24DK132733. She also receives support from the NIDDK COURAGE Program Award through the BRIDGES Consortium at Massachusetts General Hospital, the Malone Center for Engineering in Healthcare at Johns Hopkins University, and the National Institute of Minority Health and Health Disparities (3P50MD017348-03S1). Dr. Ogungbe serves as Co-Investigator on studies funded by the National Heart, Lung, and Blood Institute of the National Institutes of Health (1R01HL175642-01; OT2HL161612), the American Heart Association Health Equity Research Network (LINKED-BP), and Resolve to Save Lives, funded by Bloomberg Philanthropies, the Bill & Melinda Gates Foundation, and Gates Philanthropy Partners with support from the Chan Zuckerberg Initiative. The content is solely the responsibility of the authors and does not necessarily represent the official views of the National Institutes of Health or the American Heart Association.

## ACKNOWLEDGEMENTS

The authors extend their deepest gratitude to the participants who generously shared their time and experiences, making this research possible. We are profoundly grateful to the community members and leaders, faith community partners, and key partners who supported and guided our work, providing invaluable perspectives that shaped the recruitment approach. We also thank the THRIVE Food Is Medicine Team for their dedication throughout the study.

## ETHICS STATEMENT

This study was approved by the Johns Hopkins University School of Medicine Institutional Review Board (IRB00427492). All participants provided written informed consent in their preferred language prior to enrollment. The trial was prospectively registered at ClinicalTrials.gov (NCT06257550).

## DECLARATION OF COMPETING INTERESTS

The authors declare that they have no known competing financial interests or personal relationships that could have appeared to influence the work reported in this paper.

## DATA AVAILABILITY STATEMENT

The data that support the findings of this study are not publicly available due to participant privacy considerations, but are available from the corresponding author (O. Ogungbe) upon reasonable request.

## SUPPLEMENTAL MATERIALS

**Supplemental Figure 1:**
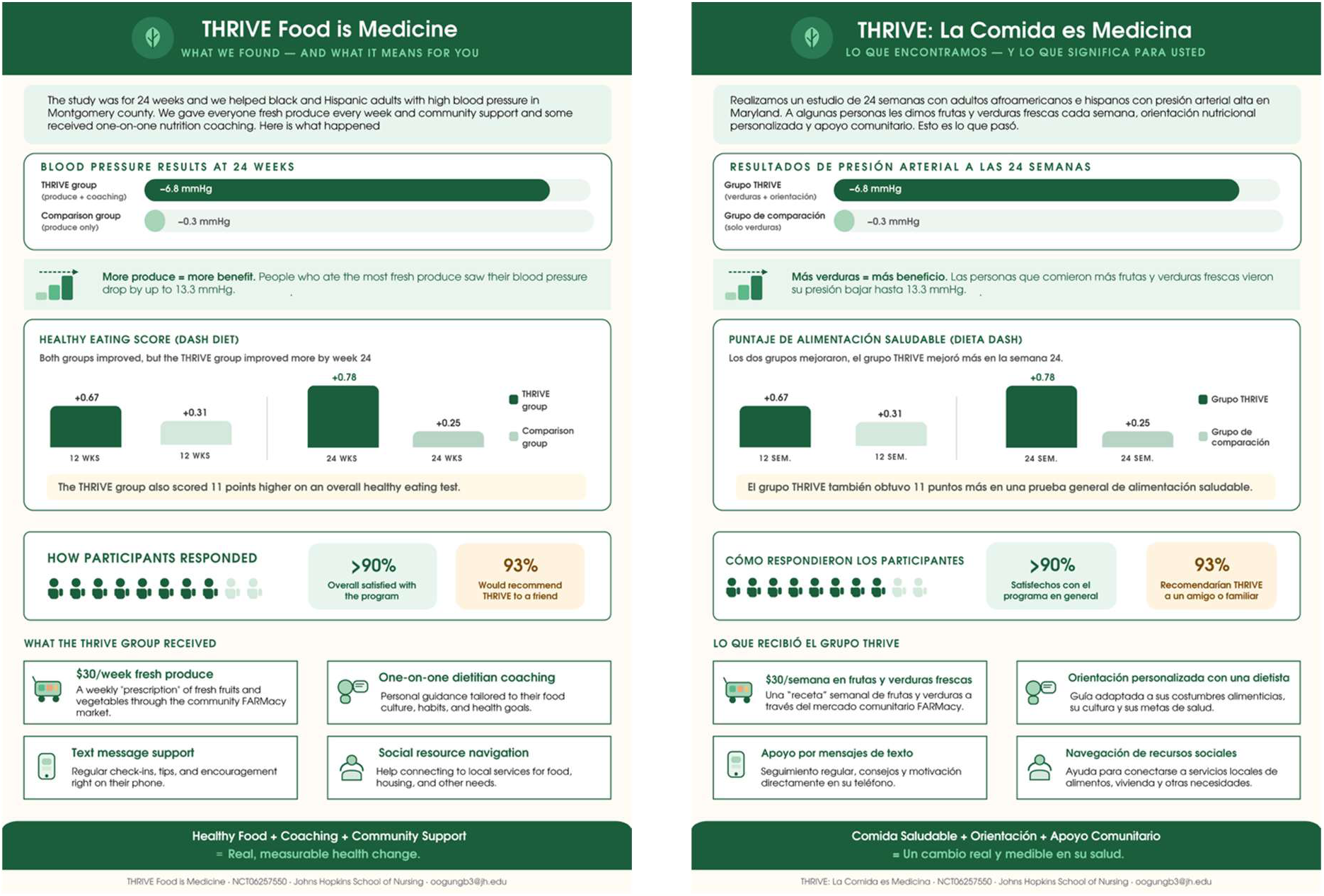
Summary of THRIVE Pilot Results Provided to Community Feedback Event Participants.

## Notes

### Competing Interest Statement

The authors have declared no competing interest.

### Clinical Trial

NCT06257550

### Author Declarations

This study was approved by the Johns Hopkins University School of Medicine Institutional Review Board (IRB00427492). All participants provided written informed consent in their preferred language prior to enrollment.

